# Rare miRNA variants in complex neuropsychiatric pedigrees alter synaptic miRNA gene-targets

**DOI:** 10.64898/2026.11.17.26367423

**Authors:** Luka Mamaladze, Cathal Ormond, Brendan Hearn, Yi-Chieh Chang, Arsalan Hassan, Muhammad Ayub, Pippa Thomson, Carol A Mathews, William Byerley, Lynn E DeLisi, Elizabeth Heron, Aiden Corvin, Niamh Ryan

## Abstract

Although rare non-coding variants are likely involved in psychiatric disorder susceptibility, their contribution is poorly understood. MicroRNAs (miRNAs) regulate gene expression, and targets in the brain include genes involved in neuronal development and synaptic plasticity. Because single-nucleotide variants (SNVs) can alter miRNA processing and target specificity, this can have disproportionate downstream impact on multiple genes affecting pathways that regulate biological processes, making them compelling candidates for mediating psychiatric disorder risk. We investigated rare miRNA variants in whole-genome sequencing (WGS) data from 25 pedigrees with diagnoses including schizophrenia, psychosis, bipolar disorder, depression, OCD, and substance use disorder. Evidence of segregation with schizophrenia and related psychotic disorders was identified for five rare miRNA variants (two seed-region, one mature-region, and two precursor-region variants), including a SNV in the mature-region of miR-4794, segregating strongly in two unrelated pedigrees and identified in a parent-offspring pair with schizophrenia from a third pedigree. One of these two pedigrees also segregates a seed-region SNV in miR-4423-3p, predicted to significantly impact expression-regulation, causing the loss of synaptic gene-targets associated with psychiatric disorders. Collectively, targets of three of the miRNAs are enriched for schizophrenia rare variant risk genes (p-value: 0.00027): six SCHEMA genes were targeted by miR-96 and four genes by miR-4423-3p.

While further understanding of miRNA function and validation is required, this study demonstrates the value of integrating structural modelling, target-shift prediction, and pathway enrichment to interpret non-coding variants. This highlights the utility of WGS for identifying and investigating rare non-coding variation in psychiatric disorder susceptibility, supplementing exome-based approaches.

## Introduction

Psychiatric disorders including schizophrenia (SCZ), bipolar disorder (BD), and major depressive disorder (MDD), are substantially heritable (1), with most cases having a complex aetiology, involving the interplay of genetic and environmental risk (2). Genome-wide association studies (GWAS) have revealed hundreds of common genetic risk variants across psychiatric disorders, individually with small effects and collectively explaining a fraction of estimated heritability (e.g. 23% for SCZ) (3–5). Studies of rare variants have identified single nucleotide variants (SNVs) and copy number variants (CNVs) that confer larger effects on disorder risk, as either *de novo* or inherited events (6–8), but individually these are carried by a small fraction of the total patient population.

Large-scale genomic studies indicate that many psychiatric disorders share a substantial proportion of their genetic architecture, with genes harbouring both common and rare variants being associated with multiple diagnoses (9). This shared architecture implies that genetic factors have pleiotropic effects that may be mediated through their impact on neurodevelopmental and/or regulatory processes (10). Most research has focussed on protein-coding variants, as their likely functional impact is easier to interpret than non-coding variants (5,11). By contrast, the contribution of higher-level regulatory variation is relatively under-explored (6). Here we investigated the role of miRNA variation on gene regulatory mechanisms.

MicroRNAs (miRNAs) represent a major class of post-transcriptional regulators with critical roles in neuronal development, synaptic plasticity, and circuit refinement (12). miRNA genes are transcribed as primary transcripts that are processed into precursor hairpins by the microprocessor complex, followed by dicer-mediated cleavage to generate mature miRNA strands (13). These short non-coding RNAs (∼22 nucleotides on average) function by binding partially complementary sequences within target messenger RNAs (mRNAs), leading to translational repression or transcript destabilisation (Figure 1). Variants occurring within different regions of the miRNA hairpin can disrupt distinct stages of this process (14). Variation within the seed region is considered the most functionally impactful, as this can alter target recognition and binding (15). Even a single nucleotide substitution within the seed region can result in the loss of canonical targets while simultaneously creating novel target interactions, substantially altering the miRNA regulatory network. Variants elsewhere within the mature miRNA may also influence target binding affinity or strand selection without completely redefining target specificity. Outside the mature miRNA sequence variation can modify downstream gene regulation by disrupting Drosha or Dicer processing, altering secondary structure stability, or affecting miRNA abundance or half-life. Because individual miRNAs can regulate hundreds of transcripts sequence variation affecting any stage of miRNA biogenesis or function has the potential to disrupt entire gene regulatory networks (16). In the brain, miRNAs are particularly abundant and dynamically regulated, with well-established roles in neurogenesis, axon guidance, and synapse remodelling (17).

**Figure 1.**
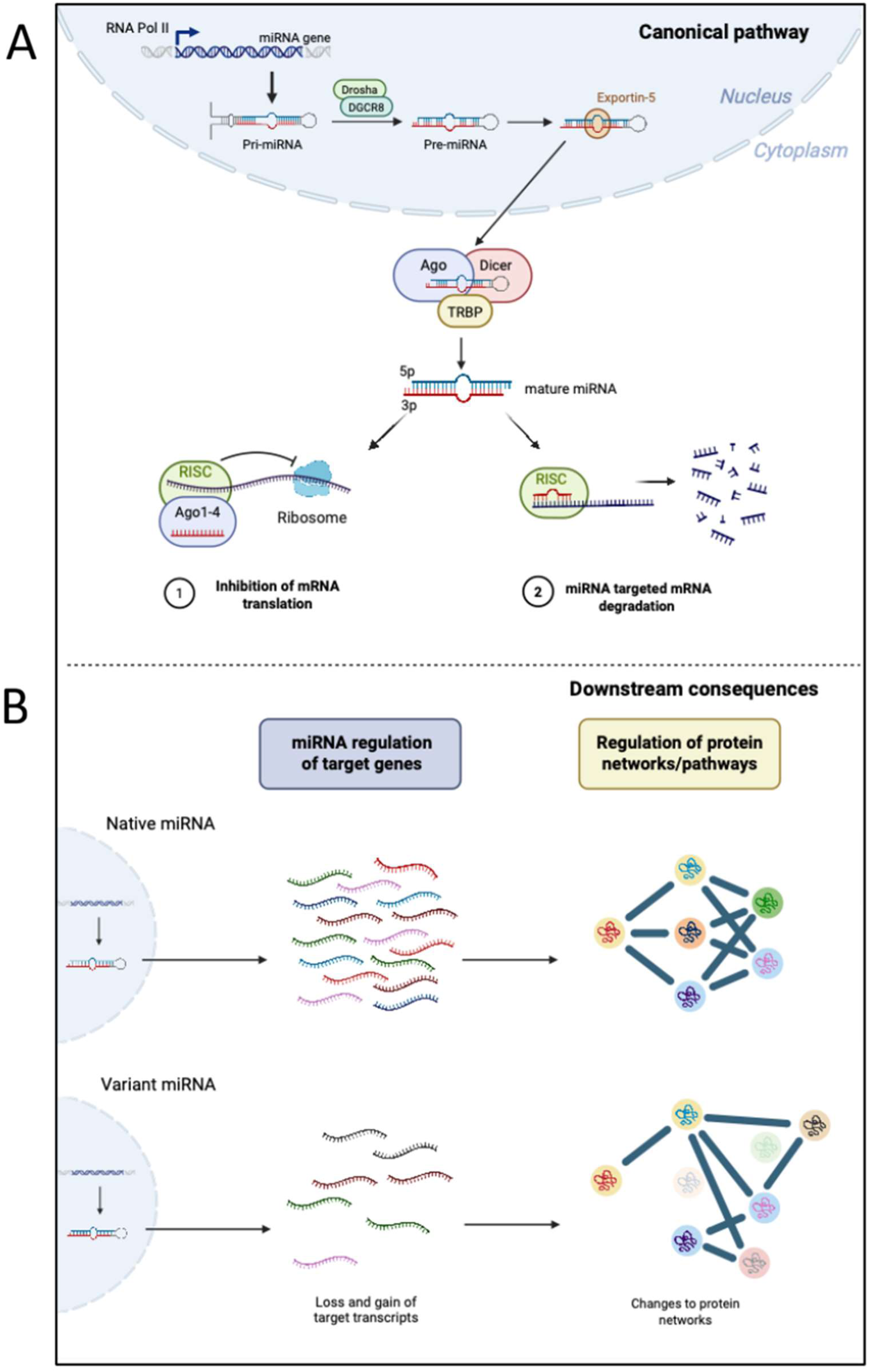
miRNA biogenesis and mechanism of action: A. Canonical pathway of miRNA biogenesis: transcription of pri-miRNA; cleavage by Drosha and DGCR8 to form pre-miRNA; transported out of nucleus and processing by DICER to form mature miRNA; B. Example of the downstream biological functions of miRNAs and potential consequences of altering miRNA sequences. Losses and gains of gene targets lead to downstream changes to protein networks and pathways. Created in BioRender. Ryan, N. (2026) https://BioRender.com/7xcddqm

Emerging data support a role for miRNAs and miRNA dysregulation in neuropsychiatric disorders including SCZ (18–20), MDD (21), BD (22), substance use disorder (SUD) (23), anxiety (24), autism (25), Tourette syndrome (26), post-traumatic stress disorder (PTSD) (27) and obsessive compulsive disorder (OCD) (28). An exemplar is miR-137, which regulates synaptic function by regulating synaptogenesis, synaptic ultrastructure and synapse function (29) and has been recognized as a crucial controller in a gene expression network implicated in neurodevelopmental processes associated with SCZ (30). miRNAs have also been shown to have the potential to be used as biomarkers (31), with miR-34a and miR-181b repeatedly being observed as overrepresented in the blood of people suffering from psychotic disorders (32). Single nucleotide polymorphisms (SNPs) in miRNA binding sites have been found to be enriched in GWAS summary statistics for SCZ, BD, MDD and anorexia nervosa, particularly for synaptic gene sets for SCZ and BD GWAS (33). SNPs in miRNA-mRNA targets in genes within dopaminergic, glutamatergic and GABAergic systems have also been associated with increased risk of SCZ (34,35).

Currently miRNA variants are poorly characterised, in part due to limitations in standard variant annotation frameworks and the lack of robust predictive tools comparable to those used for protein-coding genes (36). A recent study by Brusati et al. (2022) used an *in silico* pipeline to investigate the potential impact of rare (MAF<0.01) miRNA variants in whole genome sequencing (WGS) data from a cohort of unrelated (sporadic) amyotrophic lateral sclerosis (sALS) patients (37).

Extended pedigrees provide a powerful framework for investigating rare, high-impact genetic variation. Unlike population-based case–control designs, family-based studies enable the identification of variants that segregate with traits and allow for the assessment of inheritance patterns within a controlled genetic background (38,39). Such designs have proven valuable in identifying rare coding mutations in psychiatric and neurodevelopmental disorders (8,40–44) but have been applied less frequently to non-coding elements such as miRNA genes. Similarly, pedigree studies have been very successful at identifying rare miRNA variants and miRNA dysregulation for a number of Mendelian disorders (miR-96 and autosomal dominant deafness (45,46); miR-184 and eye diseases (47,48); miR-204 in Inherited Retinal Dystrophy (49,50); Retinitis Pigmentosa (51)) and brain disorders through altered regulation of neuronal morphology, neural induction, synaptic function, glutamatergic neurotransmission, adult neurogenesis, and neural progenitor cell proliferation (52,53).

In this study, we set out to investigate the potential contribution of rare miRNA variants to psychiatric disorders, using 25 ancestrally diverse psychiatric pedigrees. Diagnoses in these pedigrees include mood disorders, psychotic disorders, developmental and behavioural conditions, with many pedigrees having mixed diagnoses. 20 of the 25 pedigrees had previously been examined for rare coding variants, where pedigree specific deleterious missense variants were identified in genes such as *SHANK2*, *SMARCA1, NPY5R* and *ATP2B2,* but non-coding variation, including miRNA mediated regulatory variation, was not investigated (40,41,44,54,55).

## Methods

For full Methods see Supplementary Information.

### Cohort

Multiplex pedigrees were ascertained from multiple sites through the collaborative efforts of the Psychiatric Genomics Consortium (PGC) Pedigree Sequencing Working Group. The dataset consisted of 25 pedigrees comprising 202 samples from USA (mainly of European genomic ancestry) (from collaborators Bill Byerley (BB) and Lynn E. DeLisi (LDL)), Scotland (Pippa Thomson (PT)), Pakistan (Muhammad Ayub (MA)), and Costa Rica (Carol A Mathews) (Suppl. Table 1). Six samples failed sample QC filtering (three were found to be unrelated; and three due to sample-level missingness) and were excluded from analysis (final N=196). Diagnoses in these pedigrees include SCZ, psychosis, BD, MDD, OCD, attention deficit hyperactivity disorder (ADHD) and SUD. Pedigrees ranged in size from 4-16 sequenced samples.

### miRNA Variant Annotation and Filtering

See Suppl. Figure 1 for the full variant filtering pipeline. Genomic locations of human miRNAs were taken from miRBase v22 (all miRNAs excluding “Low Confidence” miRNAs; n=1,900) (56). SNVs and indels overlapping these miRNA regions were extracted and annotated by their position within the miRNA region as either precursor, mature, or seed-region miRNA variants. miRBase classifies miRNA annotations by confidence level (high, low, or not enough data). In this study, we focus on miRNAs with high-confidence annotations, while also exploring those labelled as “*not enough data*” where additional supporting evidence was identified through manual review. Population allele frequencies were incorporated using gnomAD v4.1 (57). A gnomAD-annotated VCF was generated using bcftools annotate, applying population-level allele-frequency fields from the “group max joint” metric. Rare variants were defined as having global max allele frequency < 0.01 in gnomAD v4.1.

### miRNA Segregation

Variants were prioritised based on segregation patterns within each pedigree. Variants present in all affected individuals or obligate carriers and absent from individuals unaffected by any psychiatric diagnosis were classified as highest interest (full co-segregation), followed by those present in all but one affected individual (reduced co-segregation). Where pedigrees had mixed diagnoses, this analysis focused on variants shared by the individuals with genetically-related psychiatric diagnoses, based on a recent factor analysis investigating the genetic relationship of 14 psychiatric disorders (9). Although there is known to be substantial overlap across most psychiatric disorders (see Brainstorm paper), this analysis grouped the disorders into the following five factors: Compulsive disorders (anorexia nervosa, OCD, Tourette Syndrome), Schizophrenia/Bipolar (SB) (SCZ, psychosis, BD), Neurodevelopmental disorders (autism spectrum disorder, ADHD), Internalizing disorders (PTSD, MDD, anxiety) and SUD (cannabis-use disorder, alcohol-use disorder, nicotine dependence, and opioid-use disorder). For example, in pedigree pSB285 WGS data were available for nine individuals (3 with SB, 2 with internalizing disorders (MDD) and 4 with SUD). Segregation of miRNA variants in this pedigree was assessed with respect to the SB disorders (SCZ, psychosis, BD). These diagnoses were classified as main diagnoses, whereas all other psychiatric diagnoses were classified as “other” and not considered as unaffected controls. Consequently, carriage of the candidate variant by these individuals was not required for a variant to be considered segregating within the pedigree.

To provide additional context for variant interpretation, all variants of interest identified within a pedigree were also assessed across the remaining pedigrees. Given that variant prioritisation relied heavily on segregation patterns within families and focused on rare variants, carrier status was evaluated in both affected and unaffected individuals from other pedigrees. This cross-pedigree assessment was performed to identify variants that, despite demonstrating apparent segregation within a single pedigree, may also be present in unaffected individuals from unrelated pedigrees. Such observations may reduce the likelihood that a variant represents a highly penetrant causal or risk allele. No formal exclusion thresholds were applied based on cross-pedigree carrier status; rather, these data were used to provide additional transparency and context when interpreting the strength of evidence supporting each miRNA variant.

### miRNA Target Prediction

Predicted miRNA targets were assessed for both the native sequences and, where a variant affected the seed region and thereby altered the miRNA binding sequence, the corresponding variant sequences. Firstly, mature miRNA sequences were derived from miRBase v22, and variant sequences were generated by incorporating the corresponding nucleotide substitution (58). Predicted targets for each miRNA were identified using miRDB v6.0 (59). Only targets with a stringent miRDB score greater than 80 were included (Suppl. Table 2). Gene set enrichment analysis was performed using the synapse-specific gene ontology resource SynGo v1.3 (60). SynGo performs both biological process and cellular component enrichment analysis for native and variant miRNA target sets, and pathways with a false discovery rate (FDR) adjusted p-value < 0.05 were considered significant (Suppl Tables 3-10).

### Predicted Structural Changes

Predicted structural consequences of each variant were evaluated using miRNASNP-v4 (61) and RNAfold from the ViennaRNA Package 2.0 (62), through visual assessment of the change to miRNA structure and calculating change to the Minimal Free Energy (ΔMFE), representing the amount of energy required to form its most stable secondary structure, with more negative values indicating greater structural stability.

### Overlap with known schizophrenia risk genes

A set of genes known to be associated with SCZ was generated using the SCZ whole exome sequencing (WES) study SCHEMA II (FDR < 0.05) (63) (Suppl. Table 11). An overrepresentation analysis was performed of miRNA target genes in SCHEMA II using a one-tailed Fisher’s exact test (Suppl. Methods).

## Results

### Identifying miRNA variants of interest

Across the 25 pedigrees (196 individuals), 888 variants (SNVs and indels) were identified in annotated human miRNA regions (see Supplementary Results for further details). Pedigree-level filtering identified five rare miRNA SNVs that showed inheritance patterns compatible with segregation of SB: two SNV with full co-segregation, and three with reduced co-segregation (see Table 1). Apart from miR-4794, which segregated with both SB and SUD in the same pedigree, no miRNAs were identified segregating with the other factors. Four of the variants correspond to high-confidence miRNA annotations, and one (miR-4794) occurred in a less-studied miRNA annotation (“*not enough data*”) (64). The variants mapped to different miRNA structural regions: two were located within the seed region, one in the mature miRNA sequence outside of the seed, and two in the precursor hairpin (Suppl. Figure 2). These positional differences allowed downstream comparison of how structural context relates to changes in target prediction and pathway-level consequences. Each variant was annotated (for both the native and variant miRNA) with data on potential structural changes, SynGO pathways, and whether their gene targets overlap with genes identified in rare variant association studies for SCZ.

**Table 1.** Summary of the rare miRNA variants segregating with illness in the psychiatric pedigrees, showing for each variant of interest the affected miRNA; variant ID (rs ID); location (coordinates in GRCh38); MAF: minor allele frequency (gnomAD v4.1. population max); miRNA region affected by the variant; pedigrees where the miRNA variant segregate with illness with the number of affected carriers (SB and other psychiatric diagnoses (oth)) and obligate carriers (oc). *LDL cohort ^BB cohort.

| **miRNA ID** | rsID | Coordinates | Base change | MAF | miRNA Region | Pedigree(s)<br>(carrier counts) |
|---|---|---|---|---|---|---|
| **miR-4423-3p** | rs144289108 | chr1:85133843 | T>C | 0.006 | Seed | pSB285*<br>(3/3 SB; 3/6 oth; 1 oc) |
| **miR-4749-3p** | rs147943327 | chr19:49854632 | C>T | 0.009 | Seed | K1527^<br>(4/5 SB) |
| **miR-4794** | rs191156964 | chr1:64579868 | C>T | 0.007 | Mature | K1561^<br>(5/5 SB; 1/1 oth; 1 oc)<br>pSB285 *<br>(3/3 SB; 4/5 oth; 1 oc) |
| **miR-199b** | rs146892675 | chr9:128244722 | C>T | 0.01 | Pre | K2159^<br>(3/4 SB; 1 oc) |
| **miR-96** | rs41274239 | chr7:129774734 | T>C | 0.005 | Pre | p1250*<br>(2/3 SB; 1/1 oth; 1 oc) |

### Seed region variants (miR-4423-3p and miR-4749-3p)

In pedigree pSB285 from the USA, rs144289108 was identified in the seed region of miR-4423-3p (Figure 2.A; Suppl. Figure 2.A.). This variant is carried by all three SB cases in this pedigree. Investigation of the full pedigree showed that it was also carried by the two individuals with MDD (one an affected obligate carrier), one of the four SUD individuals and an unaffected obligate carrier. rs144289108 is predicted to alter the structure of the miRNA, causing a ΔMFE of +1.7 kcal/mol, a change of +5.8%, reducing RNA stability. The native miRNA has 135 predicted gene targets, 130 of which were lost for the variant miRNA, which gained 225 new targets (Suppl. Table 2). The variant is predicted to cause a loss of synaptic pathway terms, with six of eight significantly enriched SynGO cellular component terms no longer detected for the variant miRNA (Suppl. Table 3). Similarly, four of the six SynGO biological process terms were no longer significantly enriched for the variant miRNA, which gained a third, novel term (*regulation of post-synaptic membrane potential*; FDR adj. p-value: 0.0396) (Suppl. Figure 7). While none of the targets of the native miR-4423-3p are present in SCHEMA II, four of the variant targets are SCHEMA II genes (*JARID2, SCN2A, RFX3, CACNA1B)* (Table 2).

**Figure 2.**
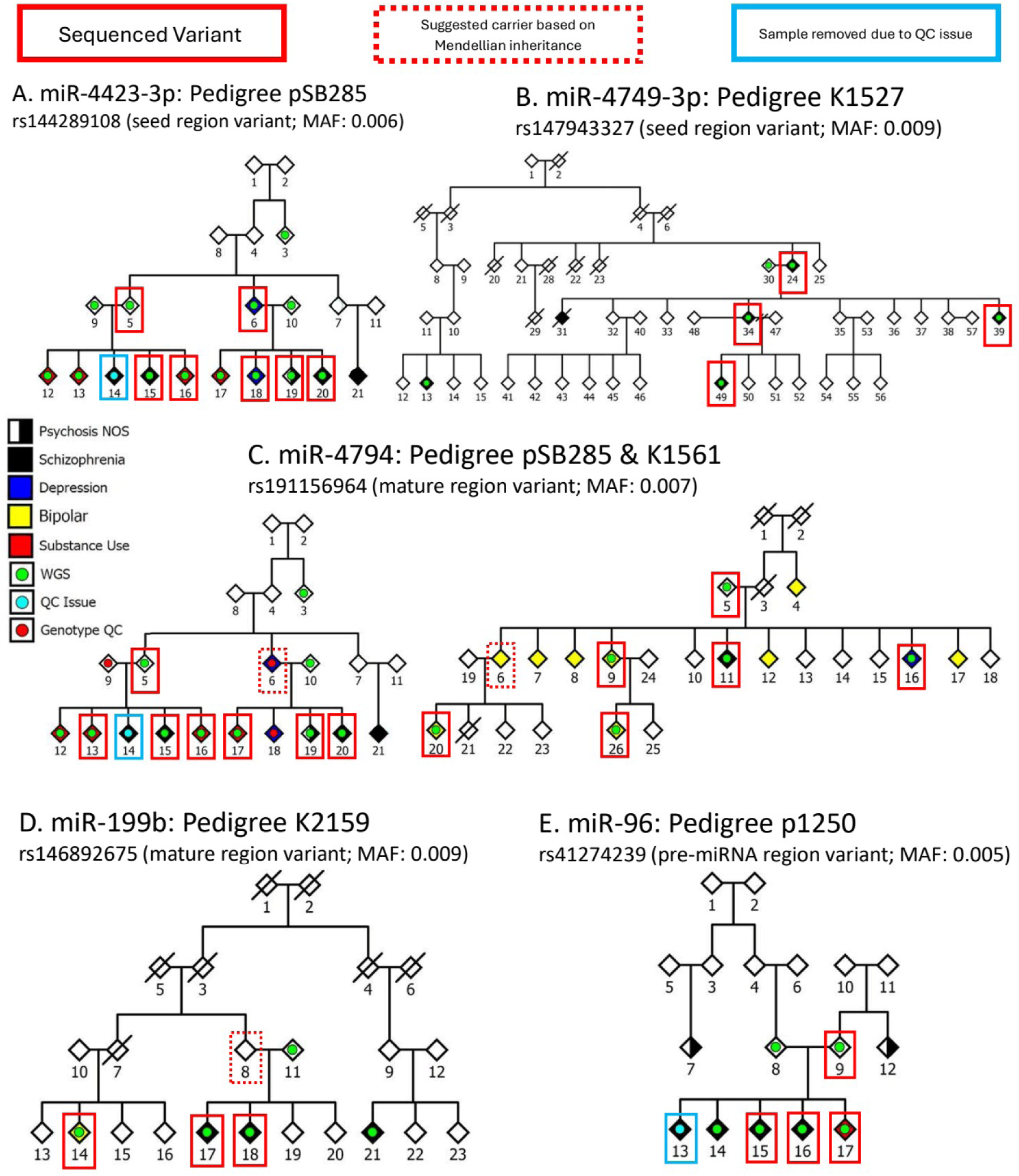
Pedigree images for families harbouring segregating rare miRNA variants. For each pedigree the individuals carrying the variant of interest are highlighted with a red rectangle; dashed rectangles indicate assumed variant carriers based on Mendelian inheritance; samples with green dots had WGS data; samples with cyan dot failed sample level QC; samples with red dot failed variant-level QC: A. miR-4423-3p variant segregating in pedigree pSB285; B. miR-4749-3p variant segregating in pedigree K1527 (BB cohort); C. miR-4794 variant segregating in two pedigrees, K1561 (BB cohort) and pSB285 (LDL cohort); D. miR-199b variants segregating in pedigree K2159 (BB cohort); and E. miR-96 variant segregating in pedigree p1250 (LDL cohort). See Supplementary Table 1 for full pedigree cohort and collaborator details.

**Table 2.**
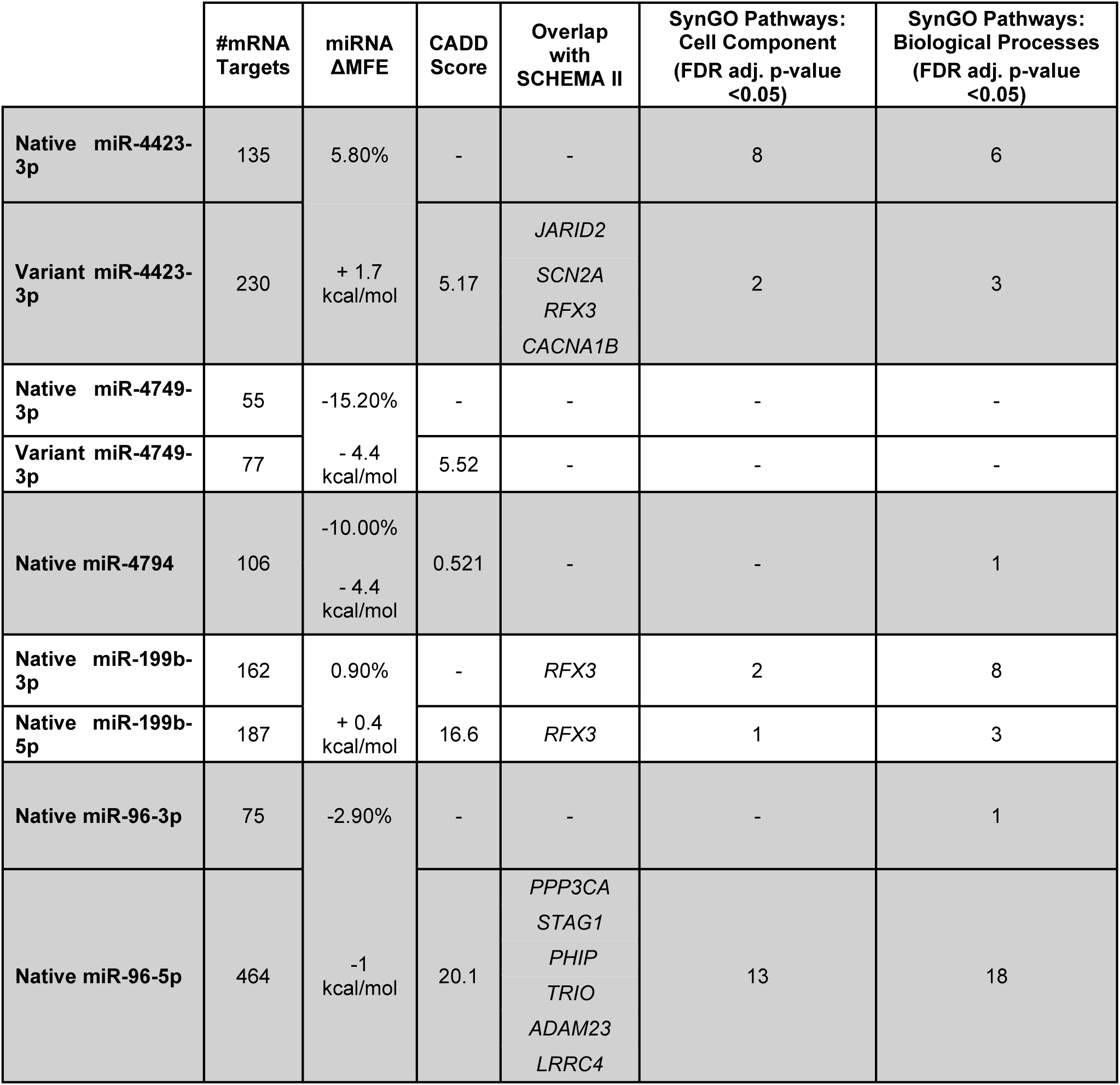
Functional annotation of the variants of interest showing for both the native and variant (when a variant was in a seed region) miRNAs: predicted number of targets; change in minimal free energy (MFE); overlap with causal genes identified in SCHEMA II; significant pathways from synapse-specific gene ontology enrichment analysis (SynGO cellular context and biological process).

|  | #mRNA Targets | miRNA ΔMFE | CADD Score | Overlap with SCHEMA II | SynGO Pathways: Cell Component (FDR adj. p-value <0.05) | SynGO Pathways: Biological Processes (FDR adj. p-value <0.05) |
|---|---|---|---|---|---|---|
| **Native miR-4423-3p** | 135 | 5.80% | - | - | 8 | 6 |
| **Variant miR-4423-3p** | 230 | + 1.7 kcal/mol | 5.17 | *JARID2*<br>*SCN2A*<br>*RFX3*<br>*CACNA1B* | 2 | 3 |
| **Native miR-4749-3p** | 55 | -15.20% | - | - | - | - |
| **Variant miR-4749-3p** | 77 | - 4.4 kcal/mol | 5.52 | - | - | - |
| **Native miR-4794** | 106 | -10.00%<br>- 4.4 kcal/mol | 0.521 | - | - | 1 |
| **Native miR-199b-3p** | 162 | 0.90% | - | *RFX3* | 2 | 8 |
| **Native miR-199b-5p** | 187 | + 0.4 kcal/mol | 16.6 | *RFX3* | 1 | 3 |
| **Native miR-96-3p** | 75 | -2.90% | - | - | - | 1 |
| **Native miR-96-5p** | 464 | -1 kcal/mol | 20.1 | *PPP3CA*<br>*STAG1*<br>*PHIP*<br>*TRIO*<br>*ADAM23*<br>*LRRC4* | 13 | 18 |

In pedigree K1527, the variant rs147943327 was identified in the seed region of miR-4749-3p (Figure 2.B; Suppl. Figure 2.D), carried by four of the five sequenced SCZ cases and absent in controls. This variant is predicted to alter the structure of the miRNA, changing both the stability (ΔMFE= –4.4 kcal/mol; a change of –15.2%), and binding capacity of the miRNA. The native miRNA is predicted to target 55 genes, while the variant miRNA gains an additional 22 targets. SynGO showed no significantly over-represented terms in either the native or variant miR-4749-3p predicted targets, possibly due to the limited number of targets (Table 2; Suppl. Table 4). This variant was also identified in three additional pedigrees in our cohort but did not pass our segregation criteria (Suppl. Figure 3). Across these three pedigrees this variant was seen in five additional cases (four SB and one MDD) and two unaffected obligate carriers (parents).

### Mature miRNA variant (miR-4794)

In pedigrees K1561 and pSB285, the variant rs191156964 was identified in the mature region of miR-4794. Across both pedigrees, the variant was carried by 8/8 SB cases (in total carried by 13 of the 14 individuals with psychiatric diagnosis in these pedigrees). In pedigree pSB285, the variant was carried by three individuals with SB, three of the four SUD individuals and one obligate carrier (Figure 2.C; Suppl. Figure 2.C). While ID_6 (MDD) has missing data for this variant, Mendelian inheritance suggests they must carry this variant. In pedigree K1561, this variant was carried by all five SB cases, one individual with MDD, and one obligate carrier (Fig 2.C). This variant was also identified in a parent-offspring pair from a second pedigree in our cohort, both of whom share a diagnosis of SCZ (Suppl. Figure 4). This variant is predicted to alters the structure of miR-4794 (Table 2), which caused a ΔMFE of –4.4 kcal/mol, a change of –10.0%, potentially increasing the stability of the miRNA (Suppl. Figure 2.C). The native miRNA has 106 predicted targets, which are enriched for a single SynGO BP pathway (*process in the synapse*, FDR adj. p-value 0.02) (Table 2; Suppl. Table 5). As this variant is outside the seed region the predicted targets remained unchanged. None of the predicted targets of miR-4794 were associated with SCZ in SCHEMA II.

### Precursor miRNA variants (miR-199b and miR-96)

In pedigree K2159, variant rs146892675 was identified in the precursor region of miR-199b (Figure 2.D; Suppl. Figure 2.B) and has a CADD score of 16.6, meaning it is predicted to be among the top 2.5% most deleterious possible alterations in the human genome (65). Three of the four SB cases in this pedigree are carriers of this variant. This variant is predicted to alter the structure of the miRNA, causing a ΔMFE of +0.4 kcal/mol (+0.9%), creating an overhang in the miRNA. As this variant is outside the seed region the predicted targets remained unchanged, however, the 3p and 5p arms of miR-199b appear to have distinct targets, which may cause structural changes that could influence strand selection or arm usage rather than target identity (Suppl. Tables 6 & 7). miR-199b-3p has 162 predicted targets and miR-199b-5p has 187 predicted targets with nine genes present in both target sets.

The predicted targets of miR-199b-3p are over-represented in two SynGO Cellular Component GO terms (*synapse, post synapse*) and for one SynGO Biological Process term (*synapse organization*) (Suppl. Figure 6). One SynGO Cellular component GO term (*post synaptic endosome*) and three SynGO Biological Process terms (*synapse organisation, synapse adhesion between pre– and post-synapse,* and *regulation of modification of postsynaptic structure*) were over-represented by the predicted targets of miR-199b-5p (Table 2; Suppl. Figure 7). Both the –3p and –5p arms are predicted to target the SCZ associated gene *RFX3* from SCHEMA II (Table 2). This variant was also identified in another individual with BD, two individuals with MDD, and an obligate carrier) from pedigree GS565 (Suppl. Figure 5).

In pedigree p1250, rs41274239 was identified in the precursor region of miR-96, a miRNA associated with autosomal dominant deafness due to dysregulation of gene regulatory networks that include synapse genes (66). This variant is carried by two of the three sequenced SCZ cases, a sibling with SUD and an unaffected obligate carrier (parent) (Figure 2.E). The variant rs41274239 is not predicted to change MFE of this miRNA, however the structure of the precursor hairpin appears to be altered (Suppl. Figure 2.E), and this variant has a CADD score of 20.1 (the highest of the five miRNA variants identified), placing it in the top 1% of deleterious variants in the human genome. The 3p and 5p arms of this miRNA have different predicted targets, miR-96-3p has 75 predicted targets while the miR-96-5p has 464 predicted targets, with an overlap in seven predicted targets. The 3p-arm predicted targets did not show an enrichment in synaptic genes, while the 5p-arm predicted targets showed eight significant pathways in the cellular component, and ten significant pathways in the biological process (Table 2; Suppl. Table 8; Suppl. Figure 8). The 5p-arm has six targets from SCHEMA II (*PPP3CA, STAG1, PHIP, TRIO, ADAM23,* and *LRRC4)*. This variant was identified in a single additional individual (no psychiatric diagnosis) from pedigree GS1283 (Suppl. Figure 6).

### Genes and GO terms altered across multiple mRNA variants

We investigated the shared FDR-significant GO terms across the five miRNAs (Supplementary Table 9). The SynGo cellular component GO term *synapse* was FDR significant for three different miRNAs in segregating in this study (miR4423-3p, miR199b-3p, and miR96-5p). *Pre-synapse* was shared between miR96-5p and miR4423-3p, and *post-synapse* was shared between miR96-5p and miR199b-3p. Two SynGo biological process GO terms were shared by four miRNAs: *process in the synapse* (miR4423-3p, miR199b-3p, miR96-5p, and miR4794) and *synapse organization* (miR4423-3p, miR199b-3p, miR199b-5p, and miR96-5p). miR96-5p shares three additional FDR-significant SynGo biological process terms with miR-199b-3p: *regulation of synapse assembly*, *regulation of post-synaptic membrane neurotransmitter receptor levels* and *process in the post-synapse*. Only one term was shared by the two miRNAs segregating in pSB285 (miR4423-3p and miR4794: *process in the synapse*).

We next compared whether there was overlap of the predicted gene targets across the identified miRNAs (both native and variant), particularly for miRNAs miR-4423-3p and miR-4794 which both have variants segregating in pSB285. Upset plots (Suppl. Figure 10) showed that while most predicted targets are unique to each miRNA, sixteen genes were shared by three miRNAs and. The predicted targets of miR-4423-3p and miR-4794 shared five genes in common: *APPL2*, *SORT1*, *FAT1*, *RSBN1L*, and *SPATA5L1* (Suppl. Table 2). Of these, *SORT1* is also a predicted target of miR96-5p.

Overall, ten genes from SCHEMA II (FDR < 0.05) were found to be targets of three of the miRNAs (miR-4423-3p, miR-96-5p, and both the 3p and 5p arms of miR-199b), eight of which are novel to SCHEMA II (*ADAM23*, *CACNA1B*, *JARID2*, *LRRC4*, *PHIP*, *PPP3CA*, *RFX3*, and *SCN2A*). Over-representation analysis showed this to be a statistically significant enrichment of SCHEMA genes in the predicted targets of the miRNAs in this study (p-value: 0.00027) (See Suppl. Fig.12).

## Discussion

Genetic variation within miRNA genes can alter miRNA biogenesis, stability, and target recognition (14). Even single-nucleotide substitutions in these regions can redirect the miRNA to entirely different targets, potentially causing substantial disruption to sensitive neural regulatory systems. Variants in miRNAs can lead to the loss of the entire miRNA, creation of new seed regions, changes to MFE, miRNA abundance or half-life, and disruption of the balance of 3p and 5p arms (14). The 3p and 5p arms usually contain sequences which target different genes, often depending on tissue type and developmental stage, which is a key evolutionary function (67). Understanding the role of miRNAs in human disease can lead to clinical benefits, such as novel biomarkers or therapeutic targets (31,68).

The aim of this analysis was to investigate rare miRNA variants segregating in psychiatric pedigrees and how they impact miRNA secondary structure and target binding. We used a pipeline modelled on the approach presented by Brusati et al. (2022) to: (i) identify rare miRNA variants from WGS data that segregate with illness in psychiatric pedigrees (Table 1, Figure 2); and (ii) predict the biological effects of these variants on miRNA folding, maturation, and target gene recognition (Suppl. Figure 2). By integrating inheritance-based filtering with functional prediction (gene set enrichment and pathway analysis), we aimed to overcome some of the limitations inherent in interpreting non-coding variation. Given the central role of miRNAs in post-transcriptional gene regulation, and the strong prior evidence implicating dysregulated miRNA networks in psychiatric disorders, we predicted that the miRNAs carrying segregating variants would be involved in biological pathways relevant to the synapse and that genes associated with psychiatric disorders are likely to be miRNA target genes. WGS followed by bioinformatic filtering and variant prioritization identified five rare miRNA variants. These all segregated with the SB diagnosis factor (SCZ, psychosis or BD) and were also found to be carried by additional family members with other psychiatric diagnoses such as MDD and SUD. No miRNA variants were identified segregating within the pedigree predominantly affected by Compulsive disorders (OCD).

Of particular note, the variant rs191156964 in miR-4794 segregated in two independent pedigrees in our cohort, pSB285 and K1561 (Figure 2.C), carried by all eight SB cases (13 of the 14 cases with any psychiatric diagnosis) and two unaffected parents (obligate carriers) across these two pedigrees. This variant was also found to be carried by a parent-offspring pair with SCZ in a third independent pedigree (Suppl. Figure 4). rs191156964 is located within the mature miRNA region of miR-4794 and is predicted to lead to a 10% increase in stability, potentially limiting miRNA abundance and raising the possibility of altered strand selection or changes to positioning in precursor miRNA machinery. miR-4794 is located in intron 2 of *CACHD1*, which has been associated with neurodevelopmental disorders (69) and is a regulator of presynaptic functions (70). Intragenic miRNAs such as miR-4794 are often co-transcribed with their host genes and thought to regulate them by both direct and indirect effects (71) *CACHD1* is a predicted target of miR-4794 (Suppl Table 2) (72,73), suggesting a potential biologically relevant function for this miRNA, warranting further investigation.

In pedigree K1527 the miR4749-3p variant rs147943327 was found to segregate in four of the five SCZ cases (absent in controls). This variant was also identified in sub-branches of three additional pedigrees, in total comprising five affected individuals (three SCZ cases, one BD and one MDD) and two unaffected parents (obligate carriers). rs147943327 is predicted to increase miRNA stability, which can also increase the miRNA half-life and affect temporal expression (74). While miR-4749 is has been shown to be expressed in the brain, especially during prenatal brain development (75) and in the central nervous system (76), SynGO analysis showed no significantly over-represented terms in either the native or variant miR-4749-3p predicted targets.

Predicting the potential biological consequence of miRNAs is limited by a number of factors, including the relative lack of knowledge around the functional consequences of variants outside of the seed region. Much like how the change to protein structure and function can be more easily predicted for missense and protein truncating variants in protein-coding genes, variants within the seed region of miRNAs can be more easily assessed than variants outside the seed region by predicting how their gene targets are altered. For example, the variant rs144289108 in miR4423-3p, segregating in pSB285, was predicted to lead to a dramatic shift in targets and regulated pathways, changing 96% of its gene targets (130 targets lost, 225 gained). These gene-level changes also led to changes in synaptic GO enrichment of the predicted target genes, with only four of the fourteen SynGO terms remaining significant for the variant form of this miRNA, along with an additional term, *regulation of post-synaptic membrane potential*. Further examination of the 225 novel targets for the variant form of miR-4423-3p by Panther GO analysis revealed a gain of statistically significant pathways (FDR < 0.05), including “*regulation of the canonical Wnt signalling pathway*” and “*regulation of neurogenesis*”. These pathways are key regulators of neurodevelopment and have been consistently associated with SCZ and BD (77,78). This suggests that re-wiring of target networks may do more than reduce native miRNA function (i.e. loss-of-function), but could also redirect regulatory activity toward novel gene sets (i.e. gain-of-function) (79) leading to altered function in the brain. Taken in conjunction with its 5.8% decreased stability, targeting of novel pathways supports both rs144289108 and miR-4423-3p as candidates for impacting risk for psychiatric disorders in this pedigree.

While studies have identified an enrichment of miRNA targets in GWAS (33) the opportunity to investigate the role of rare miRNA variants has been hindered by the fact that rare variant association studies typically rely on WES, not WGS, meaning miRNAs are excluded from the study design. However, the function of miRNAs can also be assessed through their regulation of known psychiatric risk genes. Of particular relevance is SCHEMA, which meta-analysed WES data from 24,248 schizophrenia cases and 97,322 controls to identify genes enriched for ultra-rare protein-coding variants conferring risk for schizophrenia. A recent update of the SCHEMA browser (SCHEMA II: 87,959 cases and 150,587 controls) identified 40 genes (FDR<0.05), 30 of which are novel associations not identified in SCHEMA I (6). This suggests that as the sample size increases, more genes conferring risk to schizophrenia will be identified. It is intriguing that ten of the FDR-significant SCHEMA II genes were found to be targets of three of the miRNAs investigated in this study. Over-representation analysis showed this to be a statistically significant enrichment of SCHEMA genes in the predicted targets of the miRNAs in this study (p-value: 0.00027) (See Suppl. Fig.12). Of particular note, both miR-96-5p and the variant form of miR-4423-3p are predicted to target multiple SCHEMA II genes (six and four respectively) (Table 2; Suppl. Fig.11), suggesting biological mechanisms by which these rare miRNA variants may be contributing to psychiatric aetiology in these pedigrees.

miR-96 is associated with autosomal dominant deafness (45,46), and the miR-96 precursor variant rs41274239 was found to segregate in three of the four sequenced cases in pedigree p1250 (2 SCZ; 1 SUD) (Figure 2.E). This variant has a CADD score of 20.1, placing it in the top 1% of deleterious variants in the human genome, including protein altering variants. Precursor variants are predicted to alter hairpin structure and/or stability, a region increasingly recognised as functionally sensitive (80), with potential effects on miRNA processing and abundance. This suggests the functional consequences of the miR-96 variant in this pedigree may be related to changes to the regulation of its predicted gene targets, including SCHEMA genes.

In pedigree K2159 the precursor variant rs146892675 in miR-199b, was identified in three of the four SB cases (two SCZ and one BD). This variant was also identified in three affected individuals (one BD and two MDD) in a Scottish pedigree (Suppl. Figure 5). rs146892675 has a 16.6 CADD score, placing it in the top 2.5% of deleterious variants. miR-199b has distinct 3p– and 5p-arm targeting patterns, suggesting that the variant may cause structural changes that could influence strand selection or arm usage rather than target identity. However, both arms are predicted to target the SCHEMA gene *RFX3*, also targeted by the variant form of miR-4423-3p. *RFX3* is a high confidence risk gene for ASD (https://gene.sfari.org/database/human-gene/RFX3). *De novo* and rare inherited deleterious variants in this gene have also been associated with intellectual disability and ADHD (81).

Several SCHEMA genes predicted to be targeted by these miRNAs are associated with multiple psychiatric and neurodevelopmental disorders: *TRIO* is also a neurodevelopmental disorder gene (82,83); *SLF2* (also known as *FAM178A)*, a DNA damage repair gene predicted to be a target of two of the miRNAs (miR-199b-5p and miR-96-5p), was identified in a GWAS meta-analysis of alcohol consumption in UK Biobank and AlcGen and CHARGE+ (84,85); *STAG1* is a neurodevelopmental disorder gene (OMIM 617635)(86) with additional support for SCZ from WES study of >4.5K new SCZ cases (87).

Comparing the predicted gene targets of the five miRNAs, including the variant forms, found that most genes were targeted by individual miRNAs (Suppl. Figure 9). However, as two different miRNA variants in this study segregated in pedigree pSB285 (miR4432 and miR-4794) we chose to compare their predicted gene targets and identified five genes targeted by both miRNAs (*APPL2*, *SORT1*, *FAT1*, *RSBN1L*, and *SPATA5L1*). Intriguingly, one of these genes, *SORT1*, is also a predicted target of miR96-5p, one of only sixteen genes to be targeted by three of the miRNAs from this study. While none of these miRNA’s targets are SCHEMA genes, *SORT1* is a genome-wide significant hit for general cognitive ability (meta-analysis p-value 1.32E-09) (88) and Alzheimer’s disease (meta-analysis p-value 1.13E-09 (89); meta-analysis p-value 7.5E-09 (90)).

While the convergence across variants segregating with SB was limited at the level of individual gene targets, there was greater convergence at the level of biological processes, particularly synaptic pathways. This supports a model in which genetic heterogeneity converges on shared functional systems rather than specific genes, consistent with broader findings in psychiatric genetics (91). However, as predictive methods often include false positive miRNA targets, we acknowledge that it is better to focus on shared pathways of target genes rather than specific target genes, and that functional validation would be required to confirm that *SORT1* is a target of miR-4794, miR96-5p and the variant form of miR-4423-3p.

These results, while supporting a potential role for rare miRNA variants in the major psychotic disorders (SZ, BP and psychosis), have a number of caveats that need to be acknowledged. These pedigrees are small, meaning that they are under-powered for linkage analysis. Due to the relatively under-studied nature of miRNAs, appropriate training data to generate models for predicting deleteriousness of miRNA variants is not available for statistical approaches. Rare variant association studies are also not currently designed to investigate miRNA variants, which require WGS instead of WES. Therefore, the segregation and functional prediction of these variants, while interesting, do not by themselves suggest causation. In addition, miRNA target prediction is probabilistic and requires functional validation to confirm. Individual gene targets may be erroneous, which is why gene ontology and gene set enrichment analysis are better suited to supporting functional effects of miRNAs. Despite these limitations, this study provides a unique investigation into rare miRNA variants in psychiatric disorders, taking advantage of pedigrees to identify candidate miRNAs that are suitable for further investigation. This study also highlights the value of WGS and allowing the full spectrum of genomic variants, including non-coding variation, to be investigated.

## Conclusion

We have identified five miRNA variants segregating across five unrelated psychiatric pedigrees with schizophrenia/bipolar disorder. Notably, one of these miRNA variants, in miR-4794, was found to segregate in two unrelated pedigrees (pSB285 and K1561), while pedigree pSB285 was found to segregate a second miRNA variant in miR-4423-3p. Four of the five miRNA variants were found to be carried in sub-branches of eight of the twenty-five pedigrees investigated, in total representing ten additional affected individuals, three unaffected parents (obligate carriers) and one individual with unknown diagnosis. The presence of pedigree-specific variants suggests rare, potentially high-impact events might be contributing to illness within individual families. In parallel, recurrent variants may represent more generalisable risk factors and suggest potential involvement of shared molecular mechanisms particularly underlying overlapping SZ and BD susceptibility, despite incomplete segregation in some of these pedigrees.

*In silico* prediction tools showed that these five variants exert multifaceted functional consequences, including extensive changes to gene targets, potential disruptions to secondary miRNA structure, and enrichment in synaptic gene ontology categories, many of which have previously been associated with SCZ and other psychiatric disorders (78). Significantly, FDR-significant SCHEMA II genes were found to be over-represented in the gene targets of these miRNAs.

These findings collectively implicate miRNA variation as a contributor to psychiatric disorder risk in familial contexts and highlight the importance of non-coding regulatory elements in disease genetics. This further highlights the importance of whole-genome analyses, rather than targeting protein-coding and protein altering variants as is the case for most psychiatric rare variant association studies. Although replication of these findings is necessary, there are currently insufficient publicly available datasets to enable robust independent validation. Further functional validation and expanded cohort analyses will be required to determine the extent of pathogenicity and to clarify the mechanistic impact of these variants on neuronal regulatory networks.

## Ethics

### Conflict of interest statement

The authors report no biomedical financial interests or potential competing interests.

## Author contribution

NR conceived and designed the analysis. MA, LDL, BB, PT, CAM, YC, AC collected or contributed the data. LM, CO, BH and NR performed data analyses. LM and NR wrote the paper; AC, EH, CO, CAM and LDL edited the paper. NR and AC supervised the analysis with support from EH and CO. All authors commented on drafts of the paper.

## Funding

This work was co-funded by Research Ireland [24/PATH-S/12673] and the National Institute of Health [5R01MH 124875].

## Data availability

## Notes

### Competing Interest Statement

The authors have declared no competing interest.

### Author Declarations

Joint research ethics committee of Saint James' Hospital / Tallaght University Hospital (SJH/TUH) gave full ethical approval for this work

