## Supplementary Information for "Rare miRNA variants in complex neuropsychiatric pedigrees alter synaptic miRNA gene-targets"

[1]Neuropsychiatric Genetics Research Group, Department of Psychiatry, Trinity College Dublin, Ireland, [2]Department of Psychiatry, Center for OCD, Anxiety, and Related Disorders, University of Florida, Gainesville, FL, USA, [3]University College London, UK, [4]University of Edinburgh, Edinburgh, UK, [5]Department of Psychiatry and Behavioral Sciences, University of California, San Francisco, California, USA, [6]Cambridge Health Alliance, Cambridge, MA, USA

### Supplementary Methods

#### WGS

For nine of the pedigrees (coordinated by Prof. Lynn de Lisi), details of sample ascertainment and sequencing have been published previously (1). DNA was isolated from preserved lymphoblastoid cell lines obtained from the Coriell institute for Medical Research. All samples were sequenced by deCODE Genetics with a median coverage across the coding sequence of at least 20x. For all other samples in our cohort, whole genome sequencing methods have been published previously (1,2). Briefly, DNA concentrations were quantified by Qubit or NanoDrop (Thermo Fisher Scientific), and the quality of DNA was determined by agarose gel electrophoresis. WGS was performed by Edinburgh Genomics (Clinical Genomics) on an Illumina HiSeqX to an average depth of coverage of at least 30× per sample. Some of these samples had been aligned to the GRCh38 reference genome using the Sentieon Genomics proprietary pipeline but were realigned locally to match additional data from these pedigrees.

#### Variant genotyping and QC

As described previously (2), joint genotyping was performed with GATK v3.8. Each chromosome was genotyped separately for computational efficiency, and the resulting files were combined with bcftools concat. SNVs and indels were normalised with bcftools norm. As the sample-level depth of coverage (DP) was not annotated for all variants sites, we created a new annotation (DP2) with bcftools +fill-tags representing the sum of the biallelic allele depth (AD). This new DP2 tag will always at most be equal to DP, as it represents the count of reads that were informative for genotype likelihood calculations, whereas DP represents the count of all reads at that variant site. We subsequently filtered all genotypes with genotype quality (GQ) less than 20 or with DP2 less than 10. Such genotypes were set to missing.

Variant and sample-level missingness was assessed using the plink --geno and --mind filters. Variant-level missingness was assessed first to identify poorly genotyped sites across the dataset. Variants with a missingness greater than 0.1 were removed, ensuring that only loci with genotype calls present in at least 90% of samples were retained.

Samples with variant missingness greater than 0.1 were then excluded, removing three individuals from our cohort and downstream analysis (final N= 196).

#### Over-representation analysis of miRNA targets and SCHEMA genes

SCHEMA results were obtained from the gene-based TSV files downloaded from the SCHEMA website (https://schema.broadinstitute.org/). All genes that received a p-value that were also present in GENCODE were considered as the background set (18,308). GENCODE was used to extract the gene symbol, taking v50 for Phase2.

All potential gene targets from the five prioritised miRNA variants (native and/or variant, 3p and/or 5p) were pooled (1,318 unique genes). These were then intersected with the background lists, giving a total of 1,186. SCHEMA Phase II identified 42 FDR significant (<0.05) rare variant risk genes for schizophrenia, 17 of which were exome-wide Bonferroni significant. Over-representation was evaluated using a one-tailed Fisher's exact test. Contingency tables are shown below (Suppl. Table 1). Odds ratios and their confidence intervals (CI) were calculated from the contingency tables (Suppl. Table 2, Suppl. Figure 12).

### Supplementary Results

**miRNA variant analysis**

Variants were filtered using the analysis pipeline described in Supplementary Figure 1. Across the 25 pedigrees (196 individuals), 888 variants (SNVs and indels) were identified in annotated human miRNA regions. The number of miRNA variants per pedigree ranged from 175 to 336 (155 to 297 for SNVs), reflecting differences in family size and the number of sequenced individuals. Following annotation with GnomAD v4.1 population frequencies, 238 variants met the threshold for rare variation (AF <0.01), of which 217 were SNVs. Pedigree-level filtering identified five rare miRNA SNVs that showed inheritance patterns compatible with segregation of psychiatric illness and were therefore designated as VOIs (Table 1). No indels met these pedigree-level filtering criteria. Four of the VOIs correspond to high-confidence miRNA annotations, and one (miR-4794) occurred in a less-studied miRNA annotation ("*not enough data*"). These variants corresponded to precursor, mature-miRNA, and seed-region coordinates as defined by miRBase v22.

#### Supplementary Tables and Figures:

| | | miRNA Target List | | Total |
|---|---|---|---|---|
| | | Yes | No | |
| SCHEMA II FDR <0.05 | Yes | 10 | 32 | 42 |
| | No | 1,176 | 17,090 | 18,266 |
| Total | | 1,186 | 17,112 | 18,308 |

Supplementary Table 1. Contingency table for over-representation analysis of miRNA target genes in SCHEMA II FDR significant gene set.

| SCHEMA | Odds ratio | 95% CI | p-value |
|---|---|---|---|
| Phase II | 4.54 | (1.87, 11.06) | 0.00027 |

Supplementary Table 2. Results of over-representation analysis

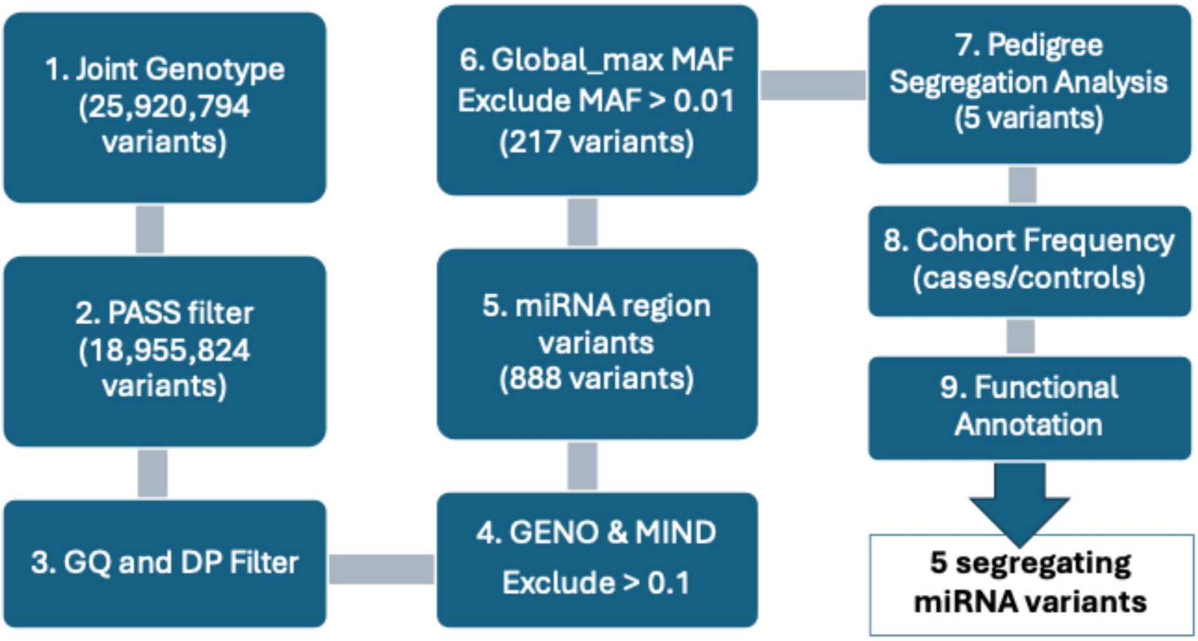

Supplementary Figure 1. Analysis Flow Chart

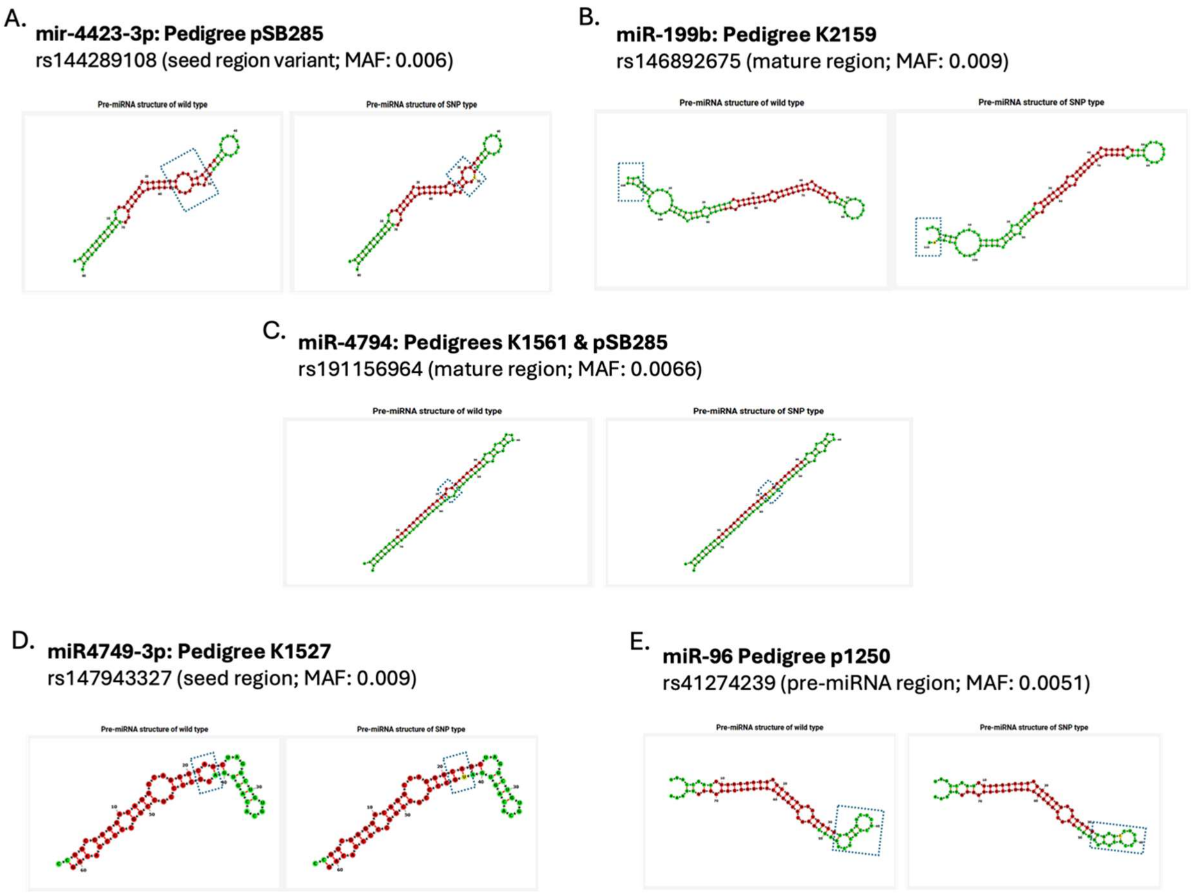

Supplementary Figure 2. Graphical representation of the segregating rare miRNA variants with their corresponding predicted miRNA structure for both the native (left structure) and variant (right structure) miRNAs (region altered by variant highlighted with navy dashed rectangle).

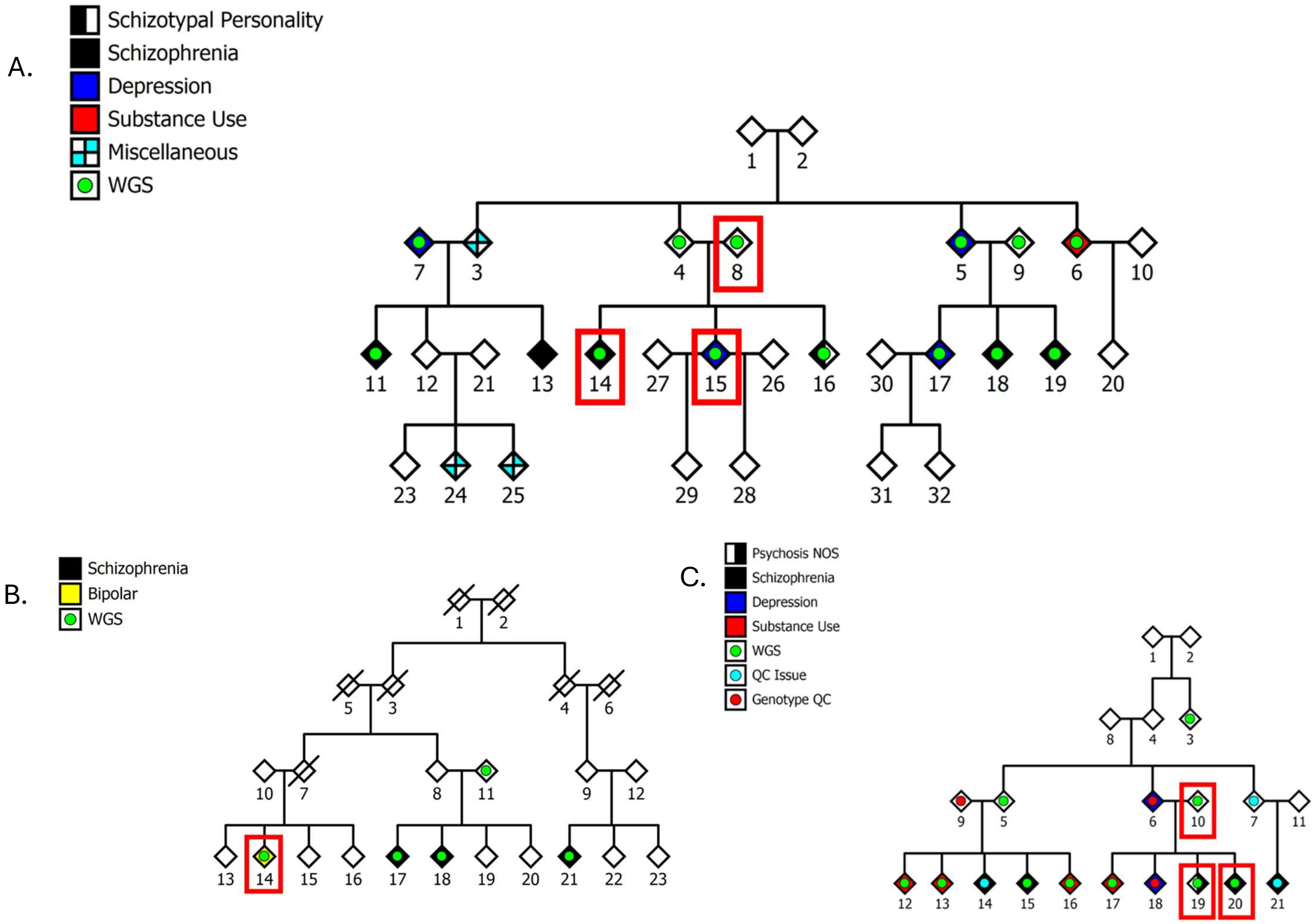

Supplementary Figure 3. Additional carriers of the miR4749-3p variant rs147943327. Individuals from three additional pedigrees (all American pedigrees of European ancestry) in the cohort were found to carry this variant, but insufficient numbers to match the strict segregation pattern reported in the main text. Additional miR4749-3p variant carriers (annotated with a cross and boxed in red) include: A. Two affected siblings (schizophrenia and depression) and unaffected parent (obligate carrier) in pedigree pVA03; B. A single affected carrier (bipolar disorder) from pedigree K2159 (parents not sequenced); C. Two affected siblings (schizophrenia and psychosis) and an unaffected parent (obligate carrier) from pedigree pSB285.

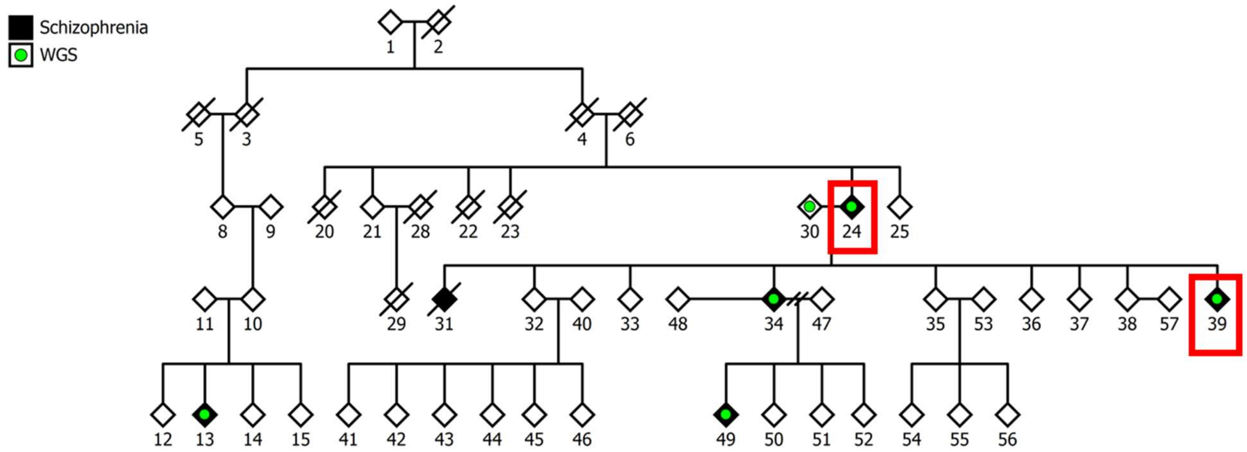

Supplementary Figure 4. Two additional carriers of the miR-4794 variant rs191156964 were identified in pedigree K1527, an American pedigree of European ancestry. These two individuals (annotated with a cross and boxed in red) are a parent-offspring pair, both affected with schizophrenia.

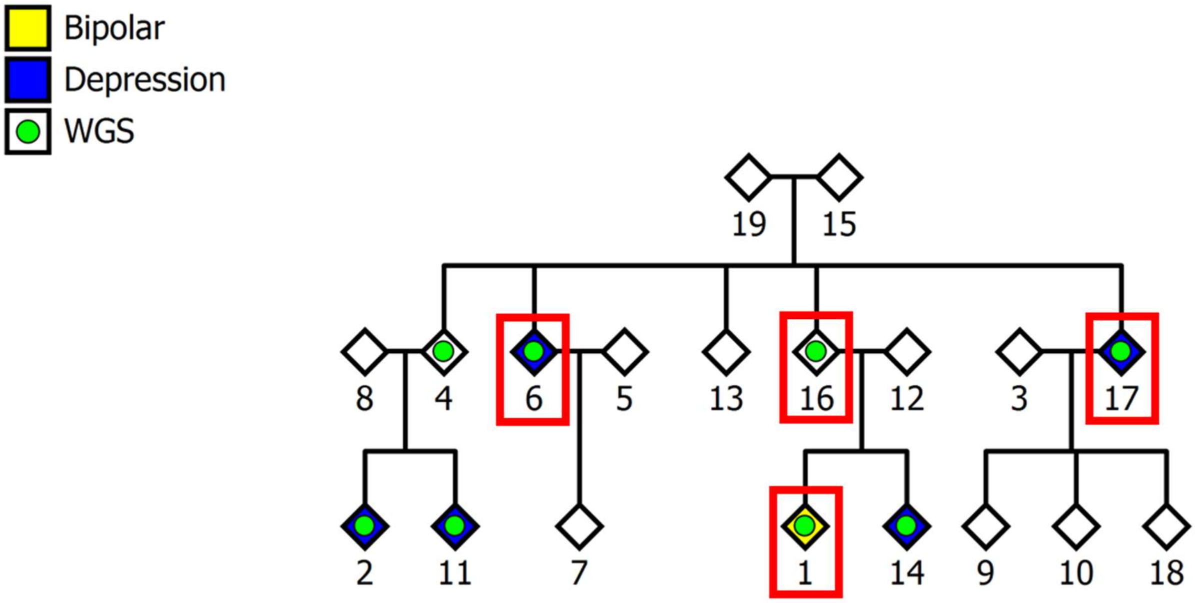

Supplementary Figure 5. Four additional carriers (annotated with a cross and boxed in red) of the miR-199b variant rs146892675 were identified in pedigree GS565, a Scottish pedigree. Three of these individuals are affected by a psychiatric disorder, two siblings with a diagnosis of depression (MDD) and their nephew having a diagnosis of bipolar disorder. The fourth carrier, while unaffected, is the parent of the BD individual, making them an obligate carrier.

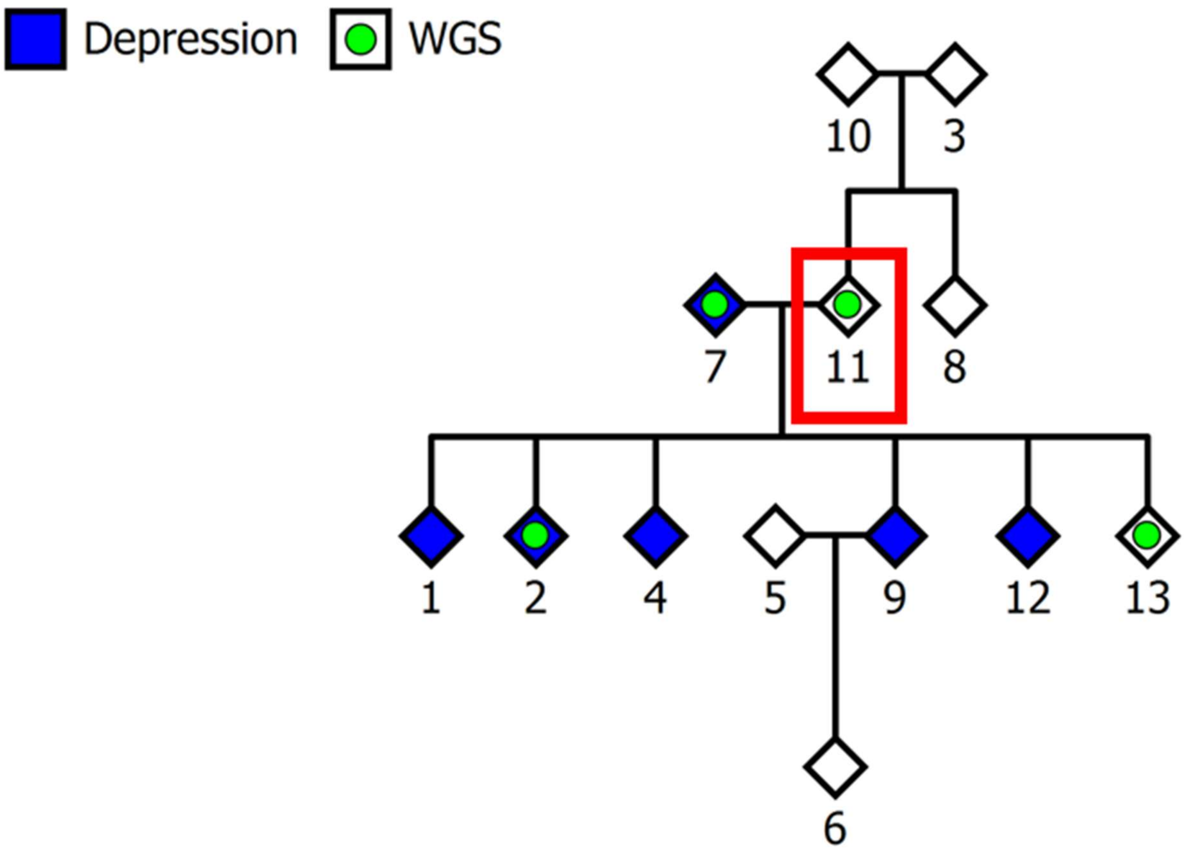

Supplementary Figure 6. An additional carrier of the miR-96 variant rs41274239 was identified in pedigree GS1283, a Scottish pedigree. It is not known whether this individual (annotated with a cross and boxed in red) has a psychiatric diagnosis. While no other members of this pedigree carry this variant it should be noted that sequencing failed for four of the five siblings diagnosed with rMDD.

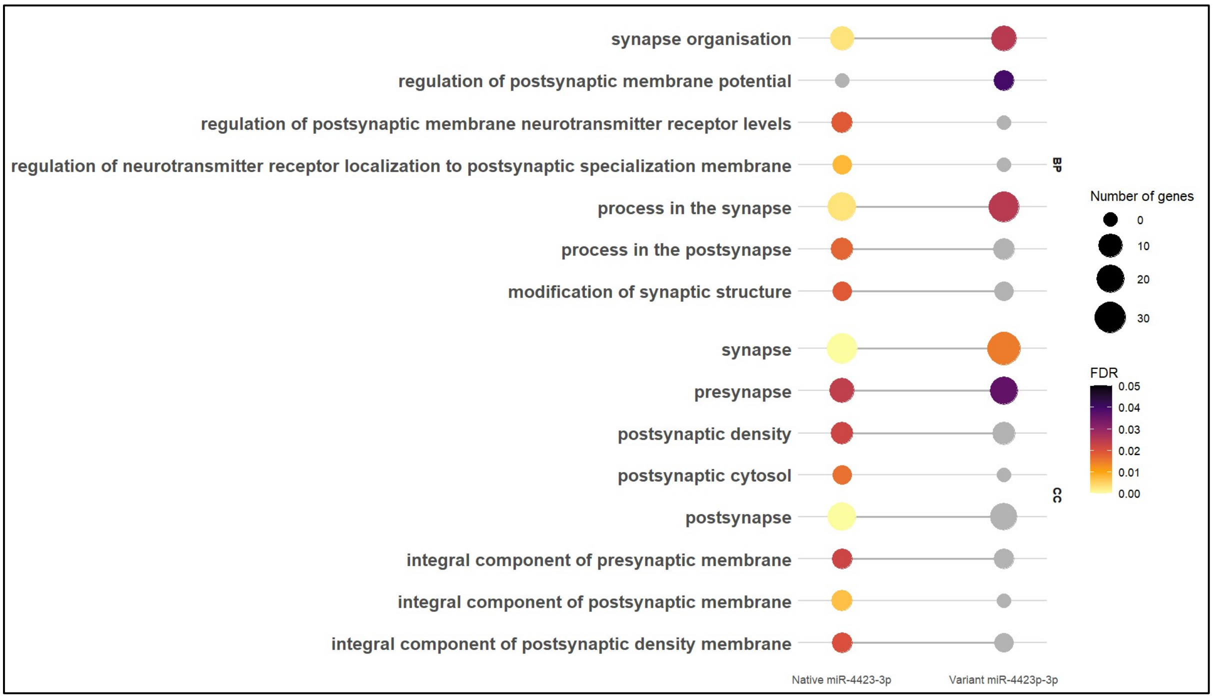

Supplementary Figure 7. SynGO gene ontology enrichment analysis for native (left dots) and variant (right dots) miR-4423-3p targets for Biological Processes (BP) and Cellular Component (CC), showing the loss of FDR significant synaptic terms for the variant miRNA targets across both CC and BP.

A.

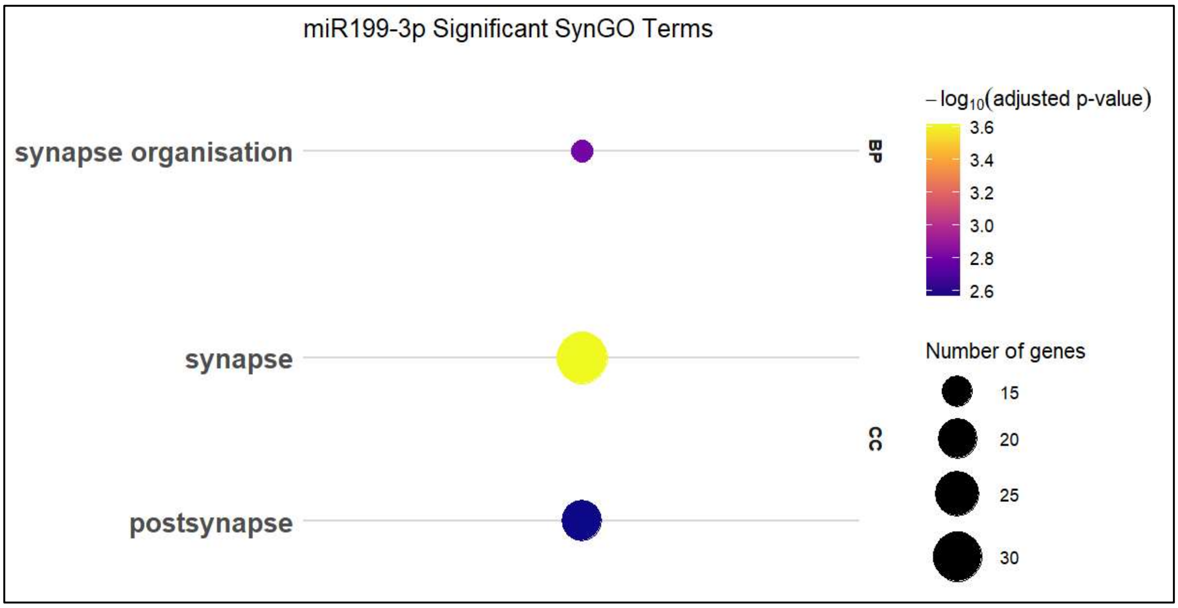

B.

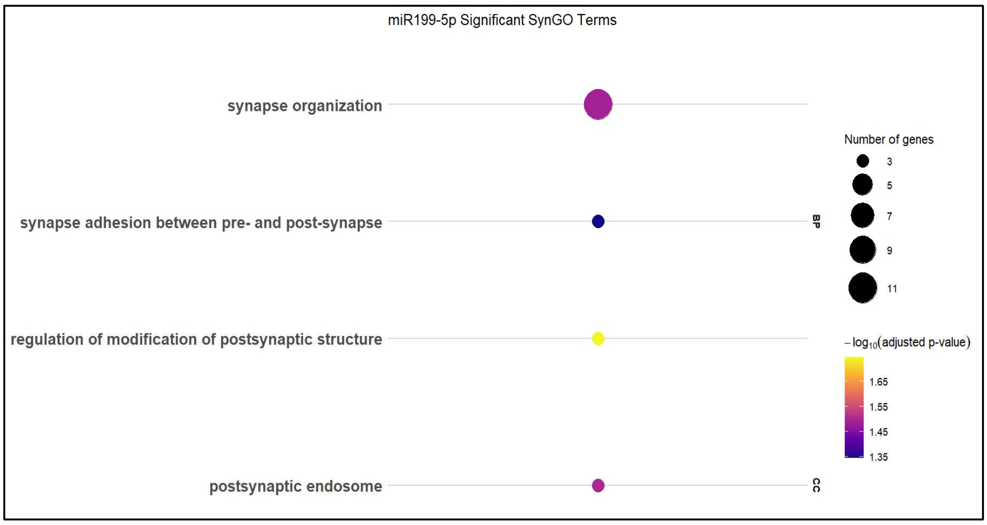

Supplementary Figure 8. SynGO gene ontology enrichment analysis for miR199b-3p (A.) and miR199b-5p (B.) targets for Biological Processes (BP) and Cellular Component (CC), showing the loss of FDR significant synaptic terms for the variant miRNA targets across both CC and BP.

A.

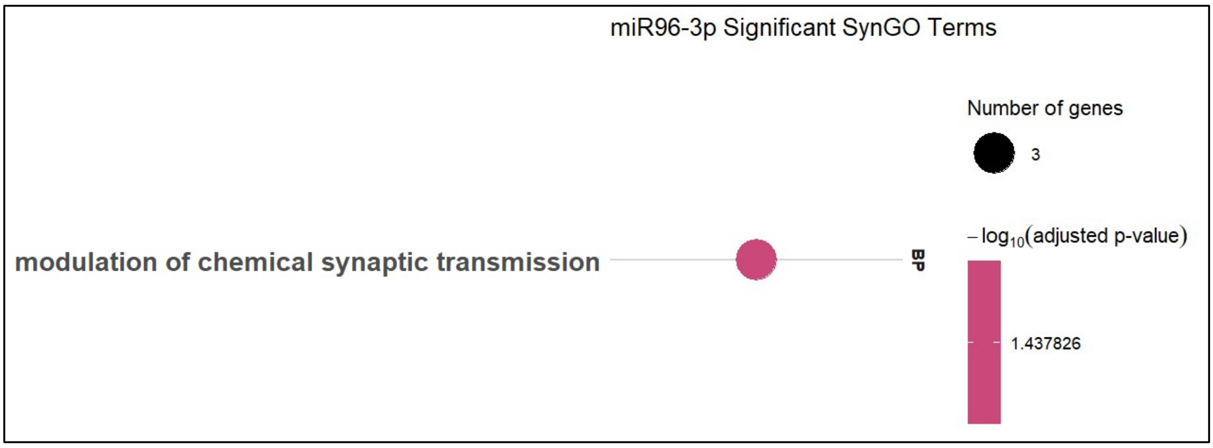

B.

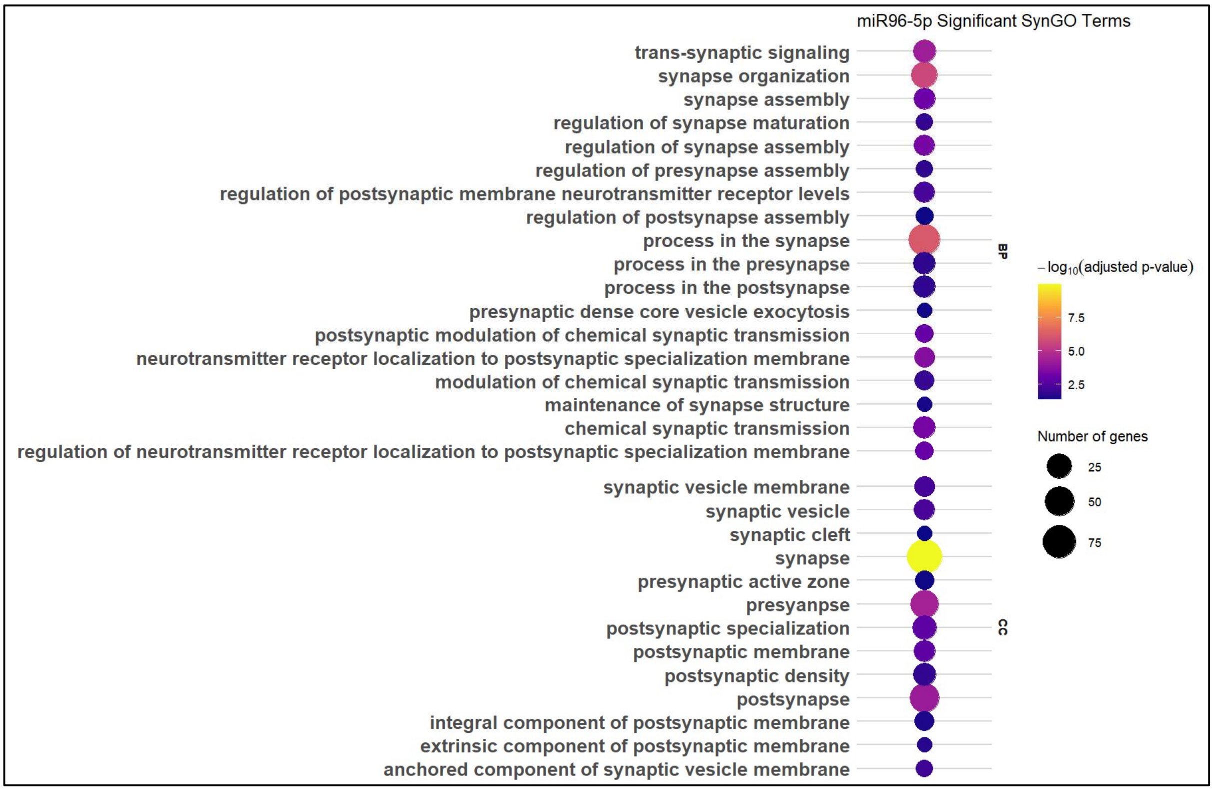

Supplementary Figure 9. SynGO gene ontology enrichment analysis for miR-96-3p (A.) and miR-96-5p (B.) targets for Biological Processes (BP) and Cellular Component (CC), showing the gain of FDR significant synaptic terms for the variant miRNA targets across both CC and BP.

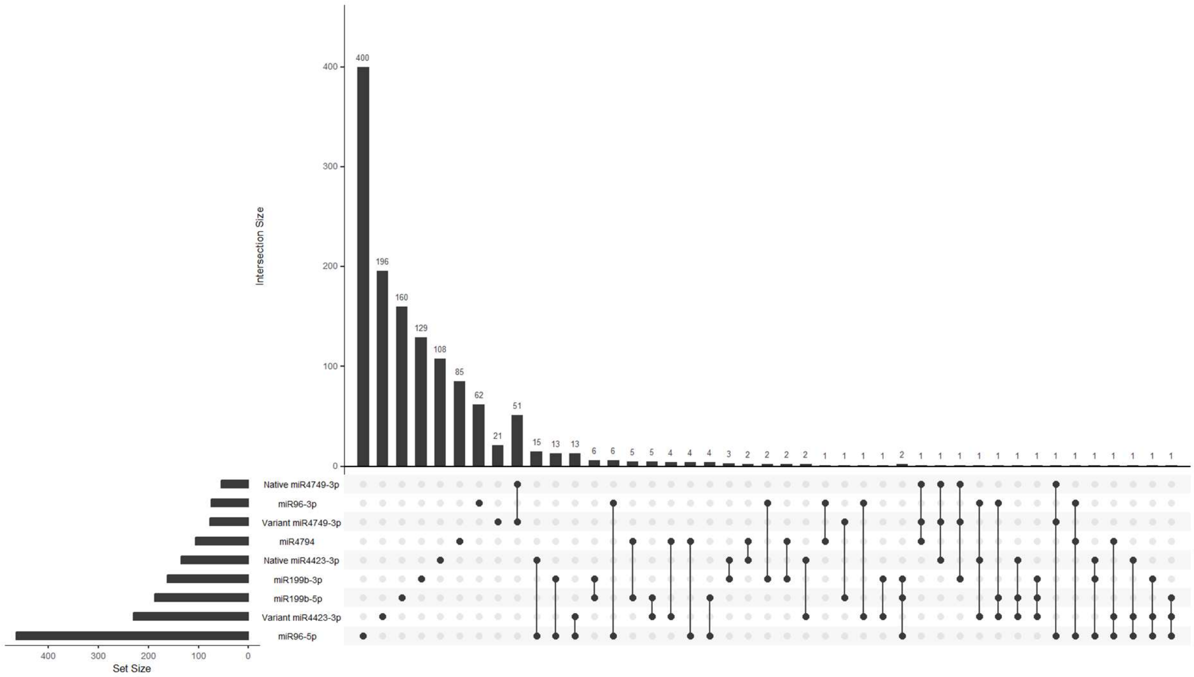

Supplementary Figure 10. Upset plot of shared vs novel targets of native and variant miRNAs. Most targets are unique to each miRNA, with a small proportion being shared by two miRNAs. No targets are common to three miRNAs.

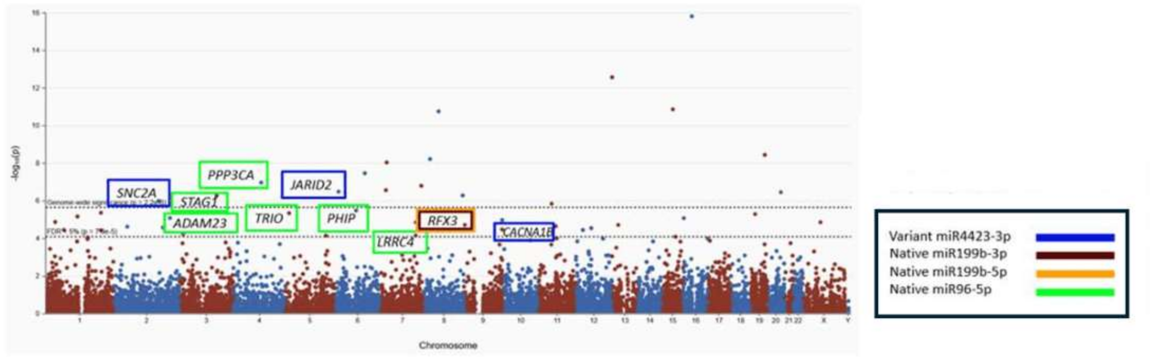

Supplementary Figure 11. Manhattan plot for SCHEMA phase II (taken from schema.broadinstitute.org); Coloured boxes highlight genes that are miRNA targets for the five miRNAs in this study (see key above).

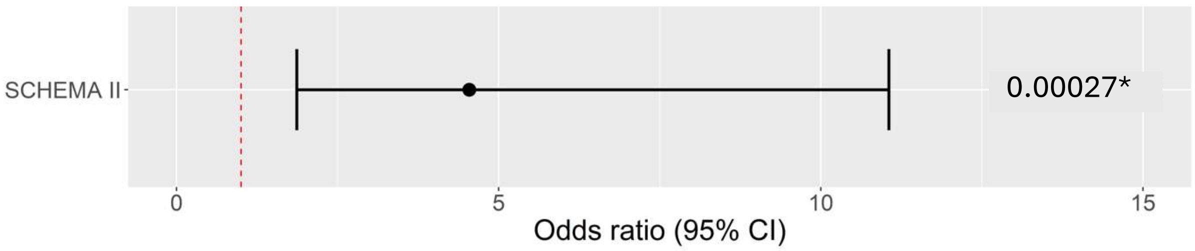

Supplementary Figure 12. Forest plot showing that SCHEMA II targets are significantly over-represented in miRNA targets following false discovery rate (FDR) adjustment.
